# Use of Semaglutide After Acute Coronary Syndrome - Design and rationale of a retrospective observational study

**DOI:** 10.1101/2025.07.31.25332452

**Authors:** M. Biasin

## Abstract

**Rationale:** Semaglutide, a glucagon like peptide 1 receptor agonist (GLP1 RA), has shown significant cardiovascular benefit in patients with type 2 diabetes mellitus (T2DM) and established atherosclerotic cardiovascular disease (ASCVD). However, its initiation in the immediate phase following an acute coronary syndrome (ACS) has not been systematically investigated. The current study aims to evaluate the early real world use of semaglutide following hospital discharge after ACS, a clinically relevant yet underexplored therapeutic window.

**Objectives:** The primary objective is to assess the real world feasibility and tolerability of semaglutide therapy, either oral or subcutaneous, when initiated at hospital discharge in patients with T2DM after ACS. Secondary objectives include the characterization of clinical and metabolic profiles of treated patients, evaluation of treatment persistence and reasons for discontinuation, documentation of adverse events, and assessment of cardiovascular outcomes during follow up.

**Methods:** This is a retrospective, observational, multicenter study including adult patients with T2DM who were hospitalized for ACS, such as ST elevation myocardial infarction (STEMI), non ST elevation myocardial infarction (NSTEMI), or unstable angina, and were discharged with a documented recommendation to initiate semaglutide. Clinical data will be extracted from electronic medical records for patients treated between January 2021 and January 2025. Collected data will include baseline demographics, ACS characteristics, cardiometabolic parameters, semaglutide formulation and dosing, as well as follow up outcomes such as treatment continuation, adverse events, laboratory parameters, and major cardiovascular events.

**Ethics and Dissemination:** This study involves retrospective analysis of anonymized clinical data. Ethical approval will be obtained in accordance with national and institutional requirements. Study results will be disseminated through peer reviewed publications and conference presentations.

**Trial Registration:** Not applicable. This is a non interventional retrospective study based on routinely collected data

## Background and Rationale

Semaglutide, a glucagon-like peptide□1 receptor agonist (GLP□1 RA), has demonstrated robust cardiovascular benefit in patients with type 2 diabetes mellitus (T2DM) and established atherosclerotic cardiovascular disease (ASCVD)^1^. However, its role in the immediate post-acute coronary syndrome (ACS) setting remains largely underexplored. To date, no studies have specifically investigated the initiation of semaglutide at the time of hospital discharge following an ACS event.

Available evidence from the PIONEER program has confirmed the cardiovascular safety and metabolic efficacy of oral semaglutide in patients with T2DM, although these trials enrolled stable outpatients rather than patients recently hospitalized for ACS ^2^. Similarly, the SOUL trial, published in 2025, demonstrated a significant reduction in major adverse cardiovascular events (MACE) with oral semaglutide in individuals with T2DM and ASCVD, but excluded patients in the immediate post-ACS phase ^3^.

Recent real-world data suggest potential benefit from early GLP-1 RA initiation after ACS. In a prospective cohort of patients with T2DM discharged after acute myocardial infarction (AMI), early combined use of GLP-1 RA and SGLT2i was associated with a significant reduction in MACE and heart failure hospitalizations compared to non-users^4^. To date, no studies have specifically addressed the initiation of GLP-1 receptor agonists at the time of hospital discharge following an ACS event, a critical window for cardiometabolic optimization that remains underutilized in clinical practice^5^.

Given the known cardiometabolic benefits of GLP-1 RAs and the emerging real-world evidence supporting their early use post-ACS, this study aims to fill a gap in the literature by retrospectively evaluating the safety, tolerability, and clinical characteristics of patients discharged from the coronary care unit (CCU) who were prescribed semaglutide (oral or injectable). By evaluating real-world use of semaglutide in the immediate post-ACS setting, this study may offer novel insights into early cardiometabolic optimization strategies in high-risk patients.

## Study Design

### Study Design

This is a retrospective, observational, multicenter study aimed at evaluating the early real-world use of semaglutide—either oral or injectable formulation—in patients with type 2 diabetes mellitus (T2DM) discharged after hospitalization for acute coronary syndrome (ACS).

The study will include adult patients admitted to the coronary care unit (CCU) with a confirmed diagnosis of ACS, including ST-elevation myocardial infarction (STEMI), non–ST-elevation myocardial infarction (NSTEMI), or unstable angina, and discharged with a documented recommendation to initiate semaglutide therapy.

Clinical data will be retrospectively collected from electronic medical records and discharge summaries of all consecutive eligible patients treated between **January 2021 and January 2025**. The following information will be extracted and analyzed: baseline demographics, details of the index ACS event, cardiometabolic profile, semaglutide formulation and dosing, and clinical outcomes during follow-up.

### Study Objectives

#### Primary Objective

- To evaluate the real-world feasibility and tolerability of semaglutide (oral or injectable) initiated at hospital discharge in patients with T2DM following an ACS event.

#### Secondary Objectives

- To describe the clinical and metabolic characteristics of patients prescribed semaglutide at discharge.
- To assess treatment persistence and reasons for discontinuation during available follow-up, regardless of predefined timepoints.
- To document adverse events potentially related to semaglutide, including gastrointestinal intolerance, hypoglycemia, pancreatitis, and renal function decline, as recorded in clinical documentation.
- To evaluate changes in key cardiometabolic parameters (HbA1c, body weight, eGFR) when available.
- To record cardiovascular outcomes during the follow-up period, including all-cause mortality, non-fatal myocardial infarction, stroke, and hospitalization for heart failure.

### Study Population

#### Inclusion Criteria

- Age ≥18 years.
- Hospital admission for ACS (STEMI, NSTEMI, or unstable angina).
- Diagnosis of type 2 diabetes mellitus (T2DM).
- Discharged from CCU with a documented recommendation to start semaglutide (oral or subcutaneous).
- Availability of medical records and at least one follow-up contact (visit or phone call).

#### Exclusion Criteria

- Terminal illness with expected survival <6 months.

### Data Collection

Data will be collected retrospectively from hospital electronic health records, discharge summaries, and outpatient clinic documentation. Follow-up data will be collected from any available clinical documentation within the observation period, including outpatient visits, phone call reports, and rehospitalizations. As this is a retrospective study, follow-up timing will not be standardized and may vary among patients. The following data will be extracted **when available**:

#### At baseline (hospital discharge)

- Demographics: age, sex, BMI.
- Cardiovascular history: previous MI, PCI/CABG, heart failure, atrial fibrillation.
- Index event details: ACS type (STEMI, NSTEMI, UA), coronary angiography findings, revascularization (PCI/CABG), LVEF.
- Laboratory: HbA1c, fasting glucose, creatinine, eGFR, lipid profile.
- Semaglutide therapy: formulation (oral/injectable), initial dose, prescriber specialty.
- Concomitant therapies: antiplatelets, statins, SGLT2i, beta-blockers, RAS inhibitors.

#### During follow-up (as available)

- Semaglutide continuation status and any dose changes.
- Reasons for discontinuation, if reported (e.g., adverse events, lack of efficacy, patient preference).
- Adverse events: GI intolerance, hypoglycemia, suspected pancreatitis, renal deterioration.
- Laboratory and anthropometric data: HbA1c, weight, eGFR.
- Cardiovascular events: death, non-fatal MI, stroke, HF hospitalization, unplanned revascularization.

## Statistical Analysis

- Descriptive statistics will be used to summarize baseline demographics, comorbidities, ACS characteristics, and treatment data. Continuous variables will be reported as mean ± SD or median [IQR], as appropriate; categorical variables will be reported as counts and percentages.
- Kaplan–Meier survival analysis will be performed to estimate time to major adverse cardiovascular events (MACE) and treatment discontinuation, with log-rank tests for group comparisons.
- Cox proportional hazards models will be used to identify independent predictors of MACE and discontinuation, adjusting for clinically relevant covariates (e.g., age, sex, eGFR, HbA1c, ACS type).
- Logistic regression will be applied for binary outcomes at specific timepoints (e.g., occurrence of adverse events at 3 or 6 months).
- Linear mixed-effects models or generalized estimating equations (GEE) will be used to analyze changes in repeated measures (HbA1c, weight, eGFR) over time, accounting for intra-subject correlation and missing data.A per-protocol sensitivity analysis will be performed to evaluate outcomes among patients who reach or maintain the target dose (14 mg/day

## Data Availability

All data in the present protocol are available upon reasonable request to the authors

